# Suicide Prevention for International Students: A Mixed Methods Evaluation of the LivingWorks safeTALK Program in Australia

**DOI:** 10.1101/2025.07.20.25331874

**Authors:** Christina Ng, Michelle Lamblin, Jo Robinson, Samuel McKay

## Abstract

International students face elevated suicide risk but are less likely to seek help than their domestic peers. This study evaluated the feasibility, acceptability, and preliminary effectiveness of an adapted version of safeTALK suicide prevention training for international students. Eight workshops were delivered in Melbourne, Australia. A total of 126 international students (60.2% female, M age = 23.4) completed surveys pre-, post-, and three months post-training, with 17 also completing follow-up interviews. The training was rated as acceptable, helpful, and safe. Linear mixed models indicated increased confidence to intervene and stronger intentions to refer individuals to formal help sources, with improvements sustained at follow-up. Suicide literacy improved, but stigma did not change, and attrition limited conclusions about long-term effects. Qualitative feedback supported the training’s value but highlighted the need for further cultural adaptation. Findings support adapted gatekeeper training as a promising strategy for suicide prevention among international students.

---

International students in tertiary education experience higher rates of suicidal behaviour compared to their domestic peers (Veresova et al., 2024). Stressors such as cultural adjustment, financial pressures, language barriers, and discrimination contribute to international students’ poor mental health and suicidality (Khanal & Gaulee, 2019; Veresova et al., 2024; Clough et al., 2018). A reluctance to seek help mental health support can further heighten their vulnerability (Clough et al., 2019). Despite this, no evidence-based suicide prevention programs are specifically designed for international students (McKay et al., 2023).

Suicide prevention approaches within education settings are typically categorised into three levels: universal, selective, and indicated (Robinson et al., 2018). Universal interventions target entire populations, such as school-wide education campaigns, selective interventions focus on at-risk subgroups, and indicated interventions address individuals exhibiting suicidal behaviours. Selective and indicated interventions can reduce suicidal thoughts and behaviours (Robinson et al., 2018). However, these types of interventions often rely on individuals actively seeking help – an area where international students face barriers due to stigma, cultural norms, and service access issues such as cost (Clough et al., 2018; McKay et al., 2023). Universal level gatekeeper training programs can overcome these barriers by equipping community members to identify and support individuals at risk without the need for active help-seeking. International student cohorts represent an ideal avenue for such universal prevention initiatives (McKay et al., 2023; Reifels et al., 2021).

Meta-analyses indicate that suicide intervention training can prevent suicide attempts and deaths in education settings (Walsh et al., 2022; Pistone et al., 2019). Training young people as “gatekeepers” enables them to support their peers while also enhancing their own mental health skills and knowledge (Pistone et al., 2019). One such intervention, Suicide Alertness for Everyone (safeTALK) training, is designed to equip community members with the skills to identify individuals at risk of suicide, understand contributing factors, and connect them with support (Burnette et al., 2015; LivingWorks Australia, 2025). safeTALK has been shown to improve suicide-related knowledge, confidence, and help-seeking behaviours, including among young people in education settings (Bailey et al., 2017; Mellanby et al., 2010). The training has also demonstrated no iatrogenic effects and is widely accepted by participants (Wilson & Neufeld, 2017). Although safeTALK has been successfully adapted for diverse groups (Mueller-Williams et al., 2023; North Western Melbourne Primary Health Network, 2022), it has not been tested with international students. Given that international students experience unique barriers to help-seeking – including cultural stigma around suicide, indirect communication norms, and lower familiarity with local mental health resources (Clough et al., 2018; McKay et al., 2023) – there is a need to explore whether safeTALK is acceptable and effective for this population.

## Current Study

This study evaluated the appropriateness, acceptability, effectiveness, and real-world application of an adapted safeTALK training for international students. We assessed if the training was engaging, relevant and culturally suitable (appropriateness and acceptability); improved suicide literacy, suicide stigma, helping self-efficacy for suicide crises, and intentions to encourage help-seeking for suicidal individuals (effectiveness); and whether participants intended to or applied the skills or knowledge in real-world situations (Application of training). The research questions were:

1. Is the training appropriate for, and acceptable to, international students?
2. Does the training change international students’ suicide prevention skills, knowledge, or stigma?
3. To what extent do participants apply the skills and knowledge gained from the training?

## Method

### Study Design

This study used a longitudinal mixed methods design. Participants provided electronic consent through a REDcap form and were assessed at three time points: Time One (T1) before training; Time 2 (T2) immediately after training; and Time 3 (T3) three months post-training. Those who completed T2 were invited to complete T3. Participants who completed T1-T3 were entered into a random draw to win one of five $100 vouchers. At T2, participants could indicate their interest in an interview and were compensated $30 for participating.

### Participants

Participants were international students aged 18 or above (M = 23.4, SD = 5.46) and enrolled in tertiary education in Victoria, Australia. Recruitment for the training was conducted through six education institutions and one student accommodation provider. A total of 126 students completed T1, 105 completed T2, and 40 students completed T3. The sample was predominantly female (60.2%), with 33.1% identifying as male and 1.5% other. Students came from diverse backgrounds, with the largest proportions from China (33.0%), Vietnam (11.28%), and India (10.5%). Regarding education level, 21.8% were enrolled in a certificate or diploma program, 31.6% in an undergraduate program, and 30.8% in a postgraduate program. Most participants reported a native language other than English (93.5%), although over half (52.6%) spoke English most of the time. Most students (91.0%) had no prior experience with suicide prevention training.

Seventeen participants (64.7% female, age M = 24.88 & SD = 6.80, from a range of countries including Vietnam, India, China, Taiwan, and Philippines) participated in an interview on Microsoft Teams conducted by CN with support from SM. Interviews lasted between 10 - 33 minutes (M = 17.58, SD = 7.74) and were audio-recorded via Microsoft Teams, automatically transcribed, and then manually reviewed.

### Intervention

LivingWorks safeTALK is a face-to-face, four-hour suicide skills alertness gatekeeper training workshop for anyone aged 15 or above. The program aims to increase trainees’ alertness to suicide and their ability to connect those thinking of suicide to further help. Eight workshops (Attendee M = 15.63, range = 9 – 32 participants) were delivered at six educational institutions and one student accommodation provider. The program was adapted for international students through a consultation workshop (See McKay et al., 2024 for a summary of the adaptation process). Adaptations emphasised the prevalence of suicidality in this population, incorporating relevant data and contextually specific examples of life stressors (e.g., academic failure, cultural shame, family and parental conflicts, racism, social status changes, and language difficulties). Barriers to discussing suicide, such as stigma, fears of burdening family, religious beliefs, language challenges, negative service experiences, and visa concerns were addressed. The program incorporated international student-specific support services and relevant role-play scenarios while retaining core safeTALK components (e.g., steps to ask about suicide, discussions, role-play activities, and video content) to maintain fidelity and key learning objectives.

### Survey Measures

#### Demographic information (T1 only)

Participants indicated their age, gender, nationality, time spent living in Australia, time spent living in other countries, education level, field of study, most spoken language, native language, training motivations and how they learned about the training.

#### RQ1: Appropriateness and Acceptability

##### Satisfaction with SafeTALK (T2 only)

Acceptability and satisfaction with the safeTALK training were assessed using a questionnaire originally developed for a school-based trial (Byrne et al., 2022). The questionnaire included five items evaluating whether participants found the training “enjoyable,” “upsetting,” and “worthwhile,” each rated on a 3-point scale (1 = not at all, 3 = very), as well as whether they would recommend the training to others (yes/no). Four additional items were developed in consultation with culturally diverse research staff to assess the cultural acceptability of the training. These items evaluated whether participants felt the training adequately prepared them to provide initial support, considered their cultural background and learning preferences, and was relevant and applicable to their cultural context (3-point scale: 1 = not at all, 3 = very).

##### Training Content Helpfulness (T2 only)

A purpose-designed questionnaire assessed the helpfulness of content areas, including suicide content, attitudes and beliefs about suicide, group simulations, role plays, and the focus on creating a “suicide-safer community”. Five items were rated on a 10-point scale (1 = Not Helpful to 10 = Very Helpful), with higher scores indicating greater perceived helpfulness.

##### General Feedback Question (T2 and T3)

Single open question asking participants if they have any further feedback about their experience of the program and/or survey.

#### RQ2: Effectiveness

##### Intentions to encourage help-seeking (T1-3)

The General Help-Seeking Questionnaire (GHSQ) assessed participants’ intentions to encourage formal and informal help-seeking for someone at risk of suicide (Wilson et al., 2005). It consists of 13 items rated on a 7-point scale (1 = Extremely Unlikely to 7 = Extremely Likely), summed separately for formal and informal sources. Higher scores indicate greater intentions to encourage help-seeking from each source. The formal (α = .90) and informal help-seeking subscales (α = .82) of the GHSQ demonstrated good internal consistency.

##### Suicide Literacy (T1-3)

Suicide literacy was assessed using the Literacy of suicide scale – short form (LOSS-SF; Batterham et al., 2013), which contains 12 items addressing common knowledge about suicide, including its causes and nature, risk factors, signs and symptoms, and treatment and prevention strategies. Items are presented in a *true, false* or *I don’t know* format. Correct responses receive a score of 1, while incorrect and *I don’t know* responses are scored 0, yielding a total score between 0 and 12.

##### Suicide Stigma (T1-3)

Suicide stigma was assessed using the Stigma of Suicide Scale - Short Form (SOSS-SF), which includes 16 items rated on a 5-point Likert scale ranging from 1 (Strongly Disagree) to 5 (Strongly Agree; Batterham et al., 2013). There are three subscales: stigma (8 items), isolation (4 items), and glorification (4 items) and scores were calculated by summing item responses, with only the stigma subscale reported in this study. Higher scores reflect greater suicide stigma. The stigma subscale demonstrated excellent reliability (α = .93).

##### Helping Self-Efficacy for Suicide Crises (T1-3)

Participants perceived self-efficacy to engage in activities to prevent or assist others in managing a suicide crisis was measured using an adapted version of the self-efficacy scale originally developed for parents of suicidal youth by Czyz et al. (2018). The adapted version included 9 items scored on a 10-point Likert scale ranging from 0 (Not Confident) to 10 (Very Confident). The items are summed, and higher scores indicate greater self-efficacy. This measure demonstrated excellent reliability (Cronbach’s α = .94).

#### RQ3: Application of Training

##### Intentions to Use Skills Learned in the Training (T2 only)

Behaviour intent to use the skills from the safeTALK training was measured with five questions asking how much participants intended to use the training and specific skills in their community on a 7-point scale ranging from 1 (Very Unlikely) to 7 (Extremely Unlikely).

##### Indication, Referral and Skill Usage (T3 only)

A purpose-designed questionnaire reflecting the core expected competencies from the safeTALK training was developed for this study. It contained 8 items assessing whether participants had used the skills they learned in the training, how they have used it and the services they referred to in the past 3 months. Questions were also asked about the outcomes of skill use or, if not used, the reasons for non-use.

### Semi-Structured Interview Guide

A semi-structured interview guided asked about participants’ perspectives on the training, including its cultural acceptability, whether they gained and applied new skills or knowledge, and suggestions for improvements. See the supplementary materials for a copy of the interview guide

### Analytic Approach

Quantitative data were analysed using RStudio (2024). Linear Mixed Models (LLM) were applied to examine the changes in the key outcome measures, including suicide literacy, suicide stigma (stigma, isolation and glorification subscale), suicide helping self-efficacy and help-seeking, over time. This model was chosen due to its ability to account for both fixed effects (e.g., time) and random effects (e.g., individual variability between participants; Hoffman, 2015).

A reflexive thematic analysis of the interview data was conducted using NVivo, following the six steps outlined by Braun and Clarke (2022). The analysis followed an interpretivist-constructivist paradigm, enabling deep exploration of students’ experiences, attitudes, and opinions while acknowledging the researcher’s reflexivity. Interview transcripts were reviewed for familiarization and analyzed inductively, with codes refined to prioritise student experiences. The codes were organised into themes, which were reviewed and refined to ensure accurate data representation and coherence in writing. CN conducted all phases of the analysis with guidance from SM.

## Results

The findings from this study are organised by research question, with quantitative results presented first, followed by qualitative insights to provide contextual depth. The qualitative analysis identified six overarching themes that capture participants’ experiences and perceptions of the training termed 1) Learning Together: An Engaging and Enjoyable Experience, 2) Accessibility and Practicality of Training Materials, 3) Cultural Relevance and Communication Styles, 4) Helping Others: Awareness and Support, 5) Readiness and Application of Skills, and 6) Empowerment and Collective Responsibility. These themes provide deeper insight into students’ experiences, complementing the statistical findings by highlighting how and why the training was effective, as well as areas for improvement.

### RQ1: Appropriateness and Acceptability

#### General Appropriateness and Acceptability

Most participants reported being “satisfied” or “very satisfied” with the training (83.8%). Nearly all participants found the training enjoyable (99.0%), worthwhile (98.1%), and not upsetting (98.1%). Additionally, 99.0% would recommend the training to others and felt it gave them the confidence to provide initial support to someone experiencing suicidal thoughts.

Participants rated the training highly (≥ 8 on a 1–10 scale), with most finding the content helpful (89.5%, M = 8.90, SD = 1.24) and valuing discussions on suicide attitudes and beliefs (85.7%, M = 8.82, SD = 1.34). Group simulations, including asking about suicide (72.4%, M = 8.45, SD = 1.73) and role plays (75.2%, M = 8.39, SD = 1.83), were well-rated. Similar ratings were received for the focus on creating a suicide-safer community (85.7%, M = 8.77, SD = 1.28). Qualitative insights from the interviews contextualize these findings by highlighting the training’s effectiveness and areas for improvement.

##### Theme 1 - Learning Together: An Engaging and Enjoyable Experience

Interview participants described safeTALK as engaging and enjoyable. This was attributed to the interactive format and skilled facilitation. They reported that the content was easy to understand, and participants appreciated that the facilitator connected well with them, making the training feel relatable and impactful. As one student said, “Generally, I find it well-structured, especially the facilitators. They were very engaging” (Participant 5).

Group discussions around cultural triggers and barriers to discussing suicide were particularly valued for providing a platform where students could share their personal experiences and learn from each other. These discussions deepened their understanding of suicide and mental health, revealing misconceptions and offering insights into different cultural approaches. “The thing I enjoyed the most was probably the discussions… it gave us an insight into… what we already know, but also some of the stuff we thought we knew but wasn’t actually true.” (Participant 10).

Students enjoyed forming bonds with others and learning about cultural similarities and differences. This allowed them to gain a more nuanced perspective and see commonalities with each other. For example, one student said they felt comfort in realising shared experiences despite coming from different cultures: “But at that moment, I feel like…, we are all human… We have the same problem sometimes” (Participant 7).

The role-playing exercises were another highlight, not just because they allowed students to practice their skills “hands-on” but also because of their interactivity: “I did like the interactive part … the questions we were asked or at the end, the little role play, I think that was like the best part for me” (Participant 17). These exercises enabled participants to apply what they had learned, which helped consolidate their knowledge: “Once I get into the role in the role-playing, I can feel I want to learn this. I want to prevent people from suicide” (Participant 2).

##### Theme 2 – Accessibility and Practicality of Training Materials

Interview participants felt the workshop was well-structured and effectively timed, appreciating the balance between presentation and interactive discussion. They appreciated the variety in the training materials, including videos, which enhanced engagement, “I love the fact they have used like different video clips to enhance the message.” (Student 9).

Timing of the training sessions was also crucial. While sessions were offered throughout the semester, students preferred having the training at the beginning when they were still adjusting to the country and facing transitional challenges: “The timing was perfect as well because it happened like in the beginning of the semester, like when students are new to the country.” (Student 8).

Participants valued the physical materials provided, such as the handbook and reminder card, which reinforced the key approaches taught in the training. However, some students found the handbook less practical outside the workshop and suggested digital versions in multiple languages for better accessibility:

I have the card with me, but the card has only so much data. So, if I had a digital version… that would be helpful for the future (Student 8).

#### Cultural appropriateness and acceptability

Most students felt that the training was designed with consideration for different cultural backgrounds (86.7%). They also believed the training was consistent with their cultural learning styles and preferences (97.1%) and found it relevant and applicable to their cultural context (86.7%). However, students also provided written feedback suggesting that it would be helpful to provide tailored approaches to conversations about mental health and suicide, including less direct phrasing, building rapport, and incorporating non-English examples to address diverse cultural norms and taboos. For instance, a student from Vietnam noted, "It’s very awkward to talk directly about mental health since it’s like a taboo topic…it could be done better by letting them feel comfortable with you first." Another participant from Singapore shared, "Asking ’Are you thinking about suicide?’ might be too confrontational…need to rephrase the question to come across less so."

##### Theme 3 - Cultural Relevance and Communication Styles

Interview participants found the language respectful and accessible, appreciating that the example scenarios were broadly applicable. One student noted that the training is not “specific to any cultural background scenario. It’s just like a normal scenario that everyone can experience.” (Participant 11).

However, some students found the direct approach challenging and suggested that adapting communication styles to align with cultural norms. For example, “The approach that you give me is a little bit direct in my opinion because, in my culture, we don’t come up to people and say, like do you think of suicide?” (Participant 11). Another said, “I feel like the skills we learn really do apply to most Australians here, but if I were to bring it back home, I don’t really think having a conversation would turn out the way we expect it to based on the training” (Participant 13).

Interview participants recommended building trust before discussing suicide, especially in family or non-Western cultural settings. For instance, a student noted, “after like having several conversations, they feel that I’m a trustworthy person. Rather than seeing them… in trouble and asking them directly” (Participant 11).

While the training demonstration videos and role-playing scenarios were helpful, they did not always resonate with interview participants’ specific experiences, who suggested including more culturally relevant scenarios to enhance relatability: “I would say it is applicable, but it lacked cross-cultural scenarios that I can apply to my family members or friends in my culture” (Participant 11). Similarly, another said, “Maybe the video examples? Like they are not like bad or anything. I just feel like they’re a little bit cliché.., I came from a very sheltered South East Asian background. We don’t talk about suicide like that.” (Participant 14). This theme highlights the need for cultural alignment in training content, but also intersects with effectiveness, as stigma and cultural barriers may limit skill application post-training and reduce reach among international students.

### RQ2: Effectiveness

#### Intentions to encourage help-seeking, Suicide Literacy, Suicide Stigma, and Self-efficacy to help someone thinking of suicide

The LMM analyses showed that intentions to encourage formal help-seeking increased significantly from T1 to T2, with these improvements largely retained at T3. In contrast, there was no significant change in intentions to encourage informal help-seeking from T1 to T2, although scores were significantly lower at T3. Suicide literacy did not increase between T1 and T2 but showed a significant improvement by T3. No significant changes were observed in suicide stigma across timepoints. Helping self-efficacy increased substantially from T1 to T2 and decreased slightly from T2 to T3, although T3 levels remained significantly higher than at baseline. A summary of the results is presented in Table 1.

**Table 1.** Raw means and standard deviations along with pairwise comparisons for help-seeking intentions, suicide literacy, suicide stigma, and helping self-efficacy across timepoints.

|  | **T1** | **T2** | **T3** | **Comparison** | **Mean Difference** | **Cohen's d** |
| --- | --- | --- | --- | --- | --- | --- |
| Formal help-seeking | 27.96<br>(6.16) | 30.98<br>(5.30) | 29.98<br>(4.48) | T1 – T2 | 2.99 [1.86, 4.13]*** | 0.70 [0.43, 0.96] |
|  |  |  |  | T1 – T3 | 2.31 [0.71, 3.91]* | 0.54 [0.16, 0.92] |
|  |  |  |  | T2 – T3 | -0.68 [-2.30, 0.94] | -0.16 [-0.54, 0.22] |
| Informal help-seeking | 20.78<br>(4.95) | 21.84<br>(4.81) | 19.18<br>(5.53) | T1 – T2 | 1.03 [-0.11, 2.16] | 0.24 [-0.03, 0.50] |
|  |  |  |  | T1 – T3 | -1.31 [-2.92, 0.29] | -0.31 [-0.68, 0.07] |
|  |  |  |  | T2 – T3 | -2.34 [-3.97, -0.71]* | -0.55 [-0.93, -0.16] |
| Suicide literacy | 8.84<br>(1.87) | 9.21<br>(1.66) | 10.12<br>(1.43) | T1 – T2 | 0.24 [–0.10, 0.58] | 0.19 [–0.08, 0.46] |
|  |  |  |  | T1 – T3 | 0.92 [0.43, 1.42]** | 0.73 [0.33, 1.12] |
|  |  |  |  | T2 – T3 | 0.68 [0.18, 1.18]* | 0.54 [0.14, 0.93] |
| Suicide stigma | 17.23<br>(7.48) | 15.67<br>(7.57) | 14.50<br>(7.43) | T1 – T2 | -1.33 [–2.58, –0.08] | -0.29 [–0.56, –0.02] |
|  |  |  |  | T1 – T3 | -1.19 [–3.15, 0.47] | -0.29 [–0.69, 0.10] |
|  |  |  |  | T2 – T3 | -0.01 [–1.83, 1.81] | 0.00 [–0.40, 0.40] |
| Helping self-efficacy | 56.62<br>(14.32) | 76.16<br>(12.28) | 69.47<br>(14.67) | T1 – T2 | 19.93 [16.99, 22.87]*** | 1.80 [1.49, 2.11] |
|  |  |  |  | T1 – T3 | 13.61 [9.39, 17.83]*** | 1.23 [0.83, 1.63] |
|  |  |  |  | T2 – T3 | -6.32 [–10.65, –1.99]* | -0.57 [–0.96, –0.18] |

Note: Values in parentheses represent standard deviations (SD). Values in square brackets represent 95% confidence intervals (CI). Cohen's d represents the standardized mean difference. \*p < .05. \*\* p < .01 \*\*\* p < .001

##### Theme 4 - Helping Others: Awareness and Support

Interview participants reported that the safeTALK training enhanced their mental health literacy and awareness of available resources. For instance, they were surprised by the number of support options available and found reassurance in knowing they were not alone, “I learned that there is more help available than I expected. I was a little bit taken aback when they showed us the slide with like all the helplines available” (Participant 14).

The training also helped normalise conversations about mental health and suicide, making students feel more confident in both supporting others and seeking help. One student mentioned, “safeTALK training let me be more sensitive to others’ emotions… I think I’ve become more proactive in caring about others and caring for myself.” (Participant 4).

Additionally, students felt that the training improved their interpersonal skills. They reported being more present and engaged in conversations, particularly when interacting with friends and peers: “The skills I’ve learnt, I think it definitely helped me in terms of being more engaged and aware in conversations when it comes to talking to friends and peers… it helps you to be more mindful.” (Participant 10).

### RQ3: Application of Training

#### Intentions to Use Skills Learned in the Training

At T2, most participants indicated (76.2%, M = 6.23, SD = 0.91) they intended to use the skills they had learned (rated as ≥ 6 on a scale from 1 to 7) particularly for raising the question of suicide (73.3%, M = 6.01, SD = 1.13), seeking more information about a plan (76.2%, M = 6.19, SD = 0.91), encouraging help-seeking (86.7%, M = 6.49, SD =0.77), calling a crisis line (81.0%, M = 6.25, SD = 0.98), and accompanying someone to get help (82.9%, M = 6.23, SD = 1.02).

#### Actual Usage of Skills Learned in the Training

At T3, 62.5% (N = 25) of participants reported using the skills gained from the safeTALK training, while 37.5% (N = 15) indicated they had not. Among those who had used the skills, 76.0% (N = 19) reported identifying invitations or signs that someone might be suicidal, and 16.0% (N = 4) stated that individuals at risk had been referred to them. Conversely, participants who had not used the skills, 86.7% (N = 13) stated that they had not interacted with someone at risk of suicide. One participant encountered someone at risk but lacked the confidence to intervene.

#### Theme 5: Readiness and Application of Skills

Interview participants generally felt well-prepared to use the skills if needed. While many had been alert to signs of distress among friends or family, few had encountered situations requiring direct intervention. For instance, one student said “Fortunately not yet I haven’t. I’ve been, yeah, looking over cues. But yeah, so far everyone around me has been doing OK” (Student 15). Participants did note that the skills could be broadly applicable, with the training empowering them to provide support to others. One recounted an experience where she assisted a friend: “Having that knowledge that I should be able to be open and understanding of the feelings of other people, I think that was a very pleasant [learning from the] workshop that helped me help my friend” (Student 12).

#### Theme 6: Empowerment and Collective Responsibility

Interview participants with lived experiences of suicide and mental ill-health found the training empowering and reassuring, appreciating the commitment of others to support those in crisis: “It also helps that knowing that people who are not experiencing these thoughts themselves are also trying to help us” (Student 14). Participating in the workshop alongside other international students fostered a sense of belonging and collective responsibility. “I think just the feeling of being involved or just included like just saying as an international student… as a collective… I think that is really good” (Student 12).

Some students were also inspired to share their newly acquired skills and knowledge with their home communities, demonstrating the training’s broader impact: “I actually do plan on … doing talks to my previous school or other schools since I think being able to just talk about that to small kids or even high school kids, they would be able to be more open about their emotions, what they’re feeling, and it would be lesser of like an awkward feeling” (Student 12).

## Discussion

This study is the first to evaluate an adapted version of safeTALK for international students. Findings indicate the training was well-received and culturally acceptable. Interactive elements, such as group discussions on cultural barriers and role-plays, were particularly valued. While cultural elements were rated highly, qualitative feedback highlighted the need for further adaptation, such as simplified language and more culturally relevant scenarios, to improve relevance. The training improved suicide literacy, self-efficacy, and formal help-seeking intentions, with many participants expressing readiness to apply these skills. By the three-month follow-up, over half of those who completed T3 surveys had applied the skills, although most interviewees had not, but felt more prepared and motivated to support others. Interviews suggested that the skills were often used in general community support rather than in crisis situations. Overall, the adapted safeTALK program appears acceptable and effective for improving key suicide prevention competencies among international students. Further cultural adaptations are warranted to enhance its reach, resonance, and long-term application. The findings align with previous evaluations of safeTALK in diverse and community samples in improving knowledge, self-efficacy and intentions to intervene (Mueller-Williams, Hopson, & Momper, 2023; Holmes et al., 2023). Similarly, Bailey et al. (2017) found that safeTALK was acceptable, safe and effective in enhancing suicide-related knowledge and confidence. These outcomes appear to stem from the program’s core components, with cultural adaptations enhancing rather than solely determining its effectiveness. A key strength of safeTALK is its structured yet interactive format, which supports its cultural adaptability. Participants particularly valued the role-play exercises, discussions on suicide attitudes and beliefs, and the focus on fostering a suicide-safer community. These findings support prior research on the value of participatory learning methods for building confidence and competence in suicide prevention training (Cross et al., 2011).

The training led to increased intentions to seek formal help and reduced reliance on informal sources, possibly reflecting a greater appreciation for professional support. Similar findings suggest gatekeeper training can shift preferences toward formal resources for international students (Kiran et al., 2023). Interviews also suggested the training fostered a sense of collective responsibility and community-mindedness. This aligns with prior research showing gatekeeper programs can empower participants to support others and take on advocacy roles by building confidence, knowledge, and willingness to intervene (Cross et al., 2011; Holmes et al., 2019).

Despite these strengths, the results point to areas for improvement. Consistent with previous studies, safeTALK did not significantly affect suicide stigma (Hashimoto et al., 2016; Mo et al., 2018). Since stigma is a significant barrier to help-seeking among international students, future adaptations of safeTALK or other interventions should consider incorporating evidence-based stigma-reduction strategies such as contact approaches. These strategies involve fostering direct or indirect interactions between individuals who hold stigmatising views and those with lived experience, aiming to reduce prejudice through personal connection and increased understanding (Patten et al., 2012). For example, sharing lived experience stories from other international students during the training could humanise the topic and challenge negative attitudes (McCullock & Scrivano, 2023).

A further challenge lies in the cultural alignment of the training. While the training was generally well-received, qualitative feedback highlighted that cultural differences in expressing distress may limit the applicability of certain skills. For instance, international students from East Asian backgrounds often rely on indirect expressions of distress, which Western-focused approaches like safeTALK – designed around direct questioning – may not adequately address (Chu et al., 2020). Without adaptations that reflect these differences, students may struggle to apply their training, reducing confidence and effectiveness (McKay & Meza, 2024). Similar challenges have been addressed in other suicide prevention programs, such as the culturally adapted QPR training in Guyana, which incorporated local risk factors, behavioural cues, and culturally appropriate rapport-building techniques (Persaud, Rosenthal, & Arora, 2019). Applying similar strategies to safeTALK could enhance its accessibility and effectiveness for international students.

International students, regardless of background, face shared stressors such as acculturation difficulties, social isolation, and challenges navigating mental health systems (Veresova et al., 2024). Rather than tailoring interventions to specific cultural groups alone, adaptations should use flexible, culturally nuanced approaches that address both diverse expressions of distress and common experiences (McKay & Meza, 2024). For safeTALK, this could include role-plays reflecting international student realities, communication strategies that accommodate direct and indirect styles, and examples of culturally relevant help-seeking. Peer-led delivery may further enhance accessibility, engagement, and cultural resonance (Muehlenkamp & Quinn-Lee, 2023; Wexler et al., 2017). However, adequate supervision and debriefing would be essential to support student facilitators and ensure fidelity (Gillard & Holley, 2018).

## Limitations

This study has several limitations. First, there was a disparity between qualitative and quantitative feedback on cultural appropriateness. Although we took steps to reduce social desirability bias, such as ensuring anonymity, conducting culturally matched interviews, and including open-ended responses, cultural norms and perceived expectations may still have led to overly favourable feedback. Future research could explore alternative methods, including implicit or behavioural measures, to better capture nuanced views. Second, participant attrition was a challenge, with only 40 participants completing the three-month follow-up compared to 105 at T2. This limits the ability to assess the sustained impact of the training and may introduce response bias, as those who remained may differ from those lost to follow-up. While reminder prompts were used, future studies should consider additional retention strategies. These could include offering incentives or using alternative follow-up formats to improve participation and strengthen conclusions about long-term effectiveness.

## Conclusion

This study provides preliminary evidence that adapted safeTALK training is acceptable and effective in improving knowledge, confidence, and intentions to intervene among international students. It increased formal help-seeking intentions but did not reduce suicide stigma, and attrition limited conclusions about long-term impact. Qualitative feedback underscored the need for further cultural adaptation. Future studies should explore culturally aligned content, peer**-**delivery models, and enhanced retention strategies. Despite limitations, findings suggest gatekeeper training holds promise as a suicide prevention approach for international students.

## Declarations

## Data Availability

The data that support the findings of this study are available upon reasonable request to the authors

## Acknowledgements

The authors would like to thank the international students who participated and LivingWorks Australia for their support of this research.

## Contributions

Christina Ng: Data curation, Formal analysis, Visualization, Writing – original draft, Writing – review & editing. Michelle Lamblin: Conceptualization, Methodology, Writing – review & editing. Jo Robinson: Conceptualization, Methodology, Resources, Writing – review & editing. Samuel McKay: Conceptualization, Funding acquisition, Methodology, Data curation, Formal analysis, Project administration, Supervision, Writing – review & editing.

## Ethics approval

The Human Research Ethics Committee at the University of Melbourne gave ethical approval for this work (No. 27010). All participants provided written informed consent before enrolment in the study, which was conducted in accordance with the Declaration of Helsinki

## Data availability statement

The data that support the findings of this study are available from the corresponding author, SM, upon reasonable request.

Clinical trial number: not applicable.

